# Development of the Aerial Remote Triage System: Result of a survey of international experts

**DOI:** 10.1101/2020.11.13.20230979

**Authors:** Cristina Álvarez-García, Sixto Cámara-Anguita, José María López-Hens, Nani Granero-Moya, María Dolores López-Franco, Inés María-Comino-Sanz, Sebastián Sanz-Martos, Pedro Luis Pancorbo-Hidalgo

**Author notes:** Corresponding authors: (NGM); (MDLF).

## Abstract

The use of drones for the triage of victims in mass-casualty incidents has recently emerged as a promising technology. However, there is not a triage system really adapted to a remote usage. The objective of our study was to develop a remote triage procedure using drones. The research was performed in three stages: literature review, development of a remote triage algorithm using drones and evaluation of the algorithm by experts. A qualitative synthesis and the calculation of content validity ratios were done to achieve the Aerial Remote Triage System. The system assesses first major bleeding, second walking, third conscious (awake) and fourth sign of life; and classify the victims inside priority categories: priority 1 (red), priority 2 (yellow), priority 3 (green) and priority * (violet). It includes the possibility to indicate save-living interventions to victims and bystanders, like the compression of bleeding injuries or the recovery position. The Aerial Remote Triage System is valid in complex health emergencies when it is difficult or impossible an immediate access to the scene due to physical, chemical or biological risks. It can be useful to know vital information about the emergencies.

## Introduction

Nowadays, the new technologies are highly important in our society, so they are increasing in the healthcare field [1]. They help to apply care faster, and it is vital in some in some life-threatening situations [2].

In recent years, the use of unmanned aerial vehicles or drones in health emergencies has increased. One of the main benefits of the use of drones is that they avoid endangering rescuers [3-6] in cases of shootings, fires, radiation or the presence of infectious agents, explosives, smoke or gases [7-8]. Drones are widely used in health emergencies because they can cover large distances in a short time and access places where rescuers had trouble reaching [9, 10], for example, rural environments [11].

One of the latest proposals is the use of drones for triage of patients in mass-causality incidents (MCIs). Some studies [4] have used the START method for triage in these accidents. The accuracy was the same when it was performed with a drone or in a standard manner, although the triage with a drone was about three and a half minutes slower, since first a triage was done using the drone so that patients who could walk left the scene, and after that a standard triage was performed. But we cannot forget that in rural areas a drone can arrive 93% faster than an emergency medical service (EMS), so it allows a remote triage before arriving at the scene of the accident. In Spain, drones have been used for an assessment of the scene before the arrival of the EMS, and it was found that on-site triage using the START method after the drone assessment was able to find 92% of the victims compared to triage without prior data that was able to find 66.7% of the victims [12]. In Canada, another study used a drone to triage with the SALT method and organize resources in a terrorist attack with explosives, and it was found that 82% of the participants adequately classified 12 of the 15 victims [13].

There are several triage systems, including START (Simple Treatment and Rapid Transport), Triage Sieve, Care Flight Triage, STM (Sacco Treatment Method), Homebush, Military Triage, CESIRA, SALT (Sort-Assess-Lifesaving Interventions-Treatment/Transport) [14]. All triage systems classify victims by a colour system, taking into account physiological parameters such as the ability to walk, breathe, perfusion/pulse or response to commands. Thus, the victims are usually classified as: (1) deceased or expectant (black), (2) immediate (red), (3) delayed (yellow), or (4) ambulatory (green). Some systems include white to classify dying patients or blue for contaminated patients [15, 16].

However, none of the existing systems are applicable in all situations, since their use is not specified, for example in situations where there may be biological, chemical, radiological or infection risks [16], poor illumination [12], rural or mountainous areas, in short in complex or difficult to access emergencies. Based on this, the objective of our study was to develop a remote triage procedure using drones.

## Materials and methods

To achieve the objective, the following phases were carried out:

### Phase 1. Literature review

The search for published studies was performed in online databases: Cuiden Plus, Global Health, LILACS, Web Of Science, Scopus, Cochrane, CINAHL, Health and Medical, IME, Medline, Science Direct and Dialnet Plus. It was not established a date limit, so the search included references from the beginning of indexing of each base until March 2020. The terms used were: unmanned aerial vehicle, drones, emergency medicine and triage. A reverse search was also carried out, so the references of the located articles that were considered of interest were analyzed.

### Phase 2. Development of a remote triage algorithm using drones

A first version of the algorithm was developed from the literature review, assessing existing triage systems. This first version evaluated the following aspects, (1) mayor bleeding, (2) walk, (3) conscious - alert, (4) sign of life. With this evaluation, patients were classified in priority 1 (red), priority 2 (yellow), priority 3 (green) and priority 4 (black). Some members of the research team presented the project in an international meeting [17], and as a result of the comments and opinions raised, it was decided to change priority 4 (black) to priority * (violet), since it is not considered ethical to classify a person as dead remotely, but should be triaged when the first responders can access the scene of an accident, thus a new colour for this condition was created, the violet (a condition between red and black). Besides, three simulations were organized to try the proposed triage system [18].

### Phase 3. Evaluation of the algorithm by experts

In order to carry out the evaluation of the algorithm, a survey was created to get expert knowledge/opinion on these statements. Professionals from different disciplines were included, academic and healthcare, from the national and international level. They were contacted by e-mail or telephone, first asking for agreeing to participate, and once they accepted, the questionnaire was sent to them. An expert panel size ranging from 10 to 15 has been shown appropriate [19]. The questionnaire was available in Spanish and English in order to enable the participation of experts of different nationalities.

The survey was implemented with SurveyMonkey platform and it was open from May 27 to June 21, 2020. The questionnaire explained clearly the instructions for filling in it and the objectives of the study.

It contained three demographic questions about respondents’ degree, work field and years of professional experience in emergencies and/or disasters. To create the second and third sections, some statements were drafted referring to the use of drones for triage. Then, two blocks of questions were delimited to facilitate the understanding: questions about the use of the drones in healthcare and, more specifically, about their use for triage where we included the proposed algorithm. Participants were asked to indicate their level of agreement with 11 consensus statements in the second section and 13 in the third section on a four-point Likert scale from 1 (completely disagree) to 4 (completely agree). Participants were encourage experts to append narrative comments to any of the statements where they were in complete or partial disagreement. Reminders were sent if the survey had not been returned. Questionnaires were coded individually. Only members of the research team had access to the codes to facilitate follow-up. Any published data identified individuals, their institution, or organizations. The IRB of University of Jaen made the exemption from ethical approval for this study, because no personal data were recorded.

A qualitative synthesis of the experts’ comments was done. Besides, we calculated content validity ratios (CVRs). We used the formula CVR = (n_e_ -N/2)/ (N/2), where n_e_ represents the number of panel experts agreeing with a statement (score of 3 or 4) and N represents the entire number of experts [20]. The optimal CVR value is set to 0.49 for a 15-experts panel [21].

## Results

The demographic characteristics of the 15 experts who agreed to participate are described in Table 1. The most of the experts were nurses (46.66%) and doctors (33.33%), they worked in academic (40%) or healthcare (40%) fields or in both (13.33%), and the mean years of professional experience was 19.53 ± 6.90.

**Table 1.**
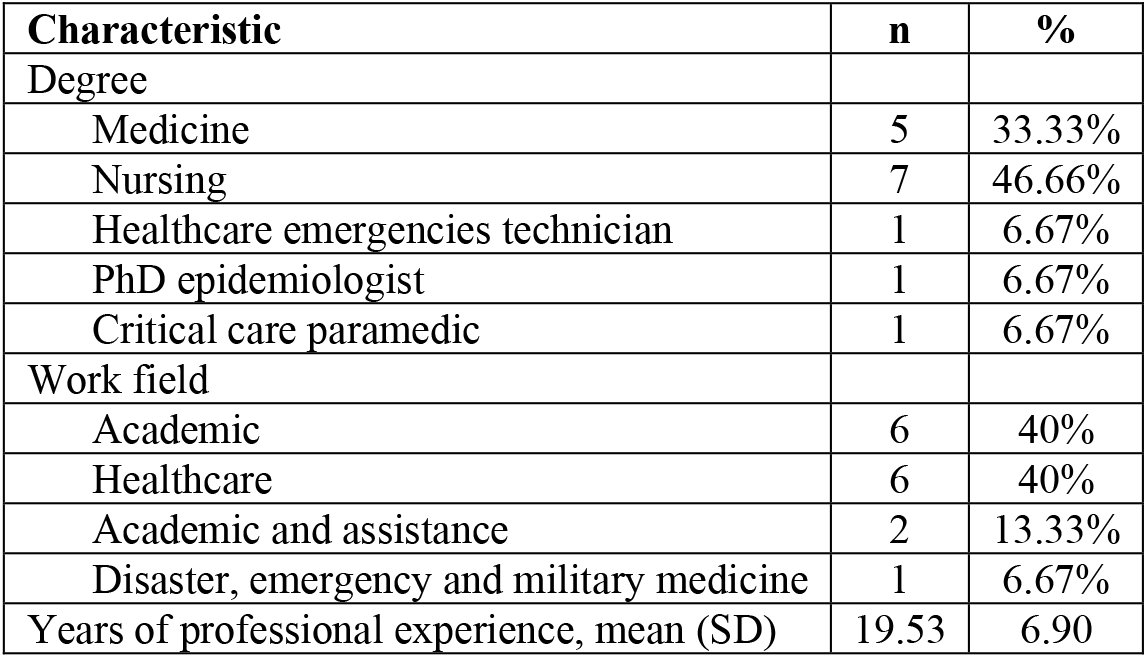
Demographic characteristics of the experts (N = 15).

| Characteristic | n | % |
| --- | --- | --- |
| Degree |  |  |
| Medicine | 5 | 33.33% |
| Nursing | 7 | 46.66% |
| Healthcare emergencies technician | 1 | 6.67% |
| PhD epidemiologist | 1 | 6.67% |
| Critical care paramedic | 1 | 6.67% |
| Work field |  |  |
| Academic | 6 | 40% |
| Healthcare | 6 | 40% |
| Academic and assistance | 2 | 13.33% |
| Disaster, emergency and military medicine | 1 | 6.67% |
| Years of professional experience, mean (SD) | 19.53 | 6.90 |

Tables 2 and 3 shows the statements along with the CVR and the average score assigned by the experts. In the block of general statements on the use of drones in disasters, only one statement did not reach the established CVR value, and in the block on specific statements on the remote triage system, two statements did not reach this value.

**Table 2.**
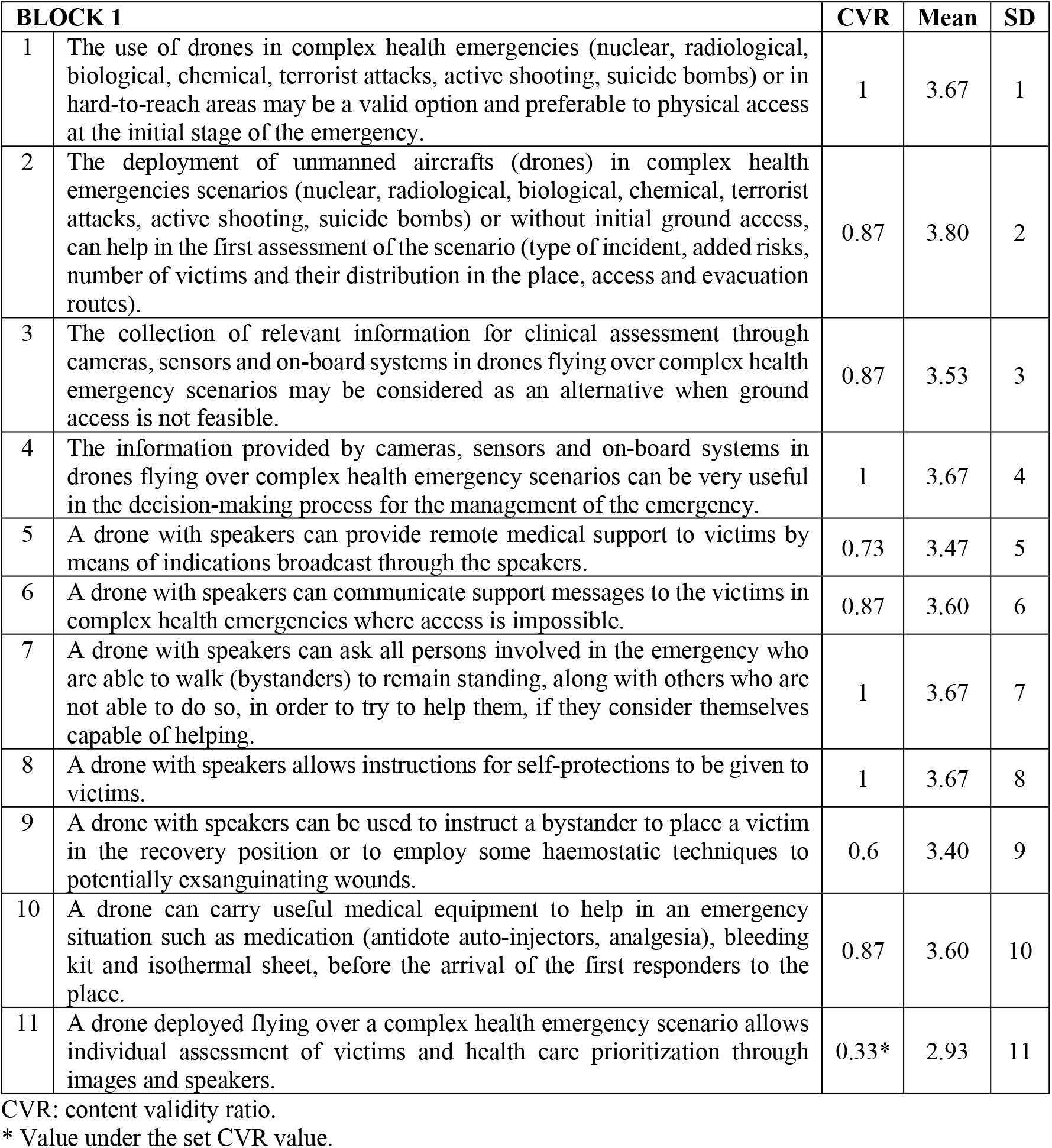
Content evaluation of statements about the use of the drones in disasters.

| **BLOCK 1** |  | **CVR** | **Mean** | **SD** |
| --- | --- | --- | --- | --- |
| 1 | The use of drones in complex health emergencies (nuclear, radiological, biological, chemical, terrorist attacks, active shooting, suicide bombs) or in hard-to-reach areas may be a valid option and preferable to physical access at the initial stage of the emergency. | 1 | 3.67 | 1 |
| 2 | The deployment of unmanned aircrafts (drones) in complex health emergencies scenarios (nuclear, radiological, biological, chemical, terrorist attacks, active shooting, suicide bombs) or without initial ground access, can help in the first assessment of the scenario (type of incident, added risks, number of victims and their distribution in the place, access and evacuation routes). | 0.87 | 3.80 | 2 |
| 3 | The collection of relevant information for clinical assessment through cameras, sensors and on-board systems in drones flying over complex health emergency scenarios may be considered as an alternative when ground access is not feasible. | 0.87 | 3.53 | 3 |
| 4 | The information provided by cameras, sensors and on-board systems in drones flying over complex health emergency scenarios can be very useful in the decision-making process for the management of the emergency. | 1 | 3.67 | 4 |
| 5 | A drone with speakers can provide remote medical support to victims by means of indications broadcast through the speakers. | 0.73 | 3.47 | 5 |
| 6 | A drone with speakers can communicate support messages to the victims in complex health emergencies where access is impossible. | 0.87 | 3.60 | 6 |
| 7 | A drone with speakers can ask all persons involved in the emergency who are able to walk (bystanders) to remain standing, along with others who are not able to do so, in order to try to help them, if they consider themselves capable of helping. | 1 | 3.67 | 7 |
| 8 | A drone with speakers allows instructions for self-protections to be given to victims. | 1 | 3.67 | 8 |
| 9 | A drone with speakers can be used to instruct a bystander to place a victim in the recovery position or to employ some haemostatic techniques to potentially exsanguinating wounds. | 0.6 | 3.40 | 9 |
| 10 | A drone can carry useful medical equipment to help in an emergency situation such as medication (antidote auto-injectors, analgesia), bleeding kit and isothermal sheet, before the arrival of the first responders to the place. | 0.87 | 3.60 | 10 |
| 11 | A drone deployed flying over a complex health emergency scenario allows individual assessment of victims and health care prioritization through images and speakers. | 0.33* | 2.93 | 11 |

CVR: content validity ratio.
\* Value under the set CVR value.

**Table 3.**
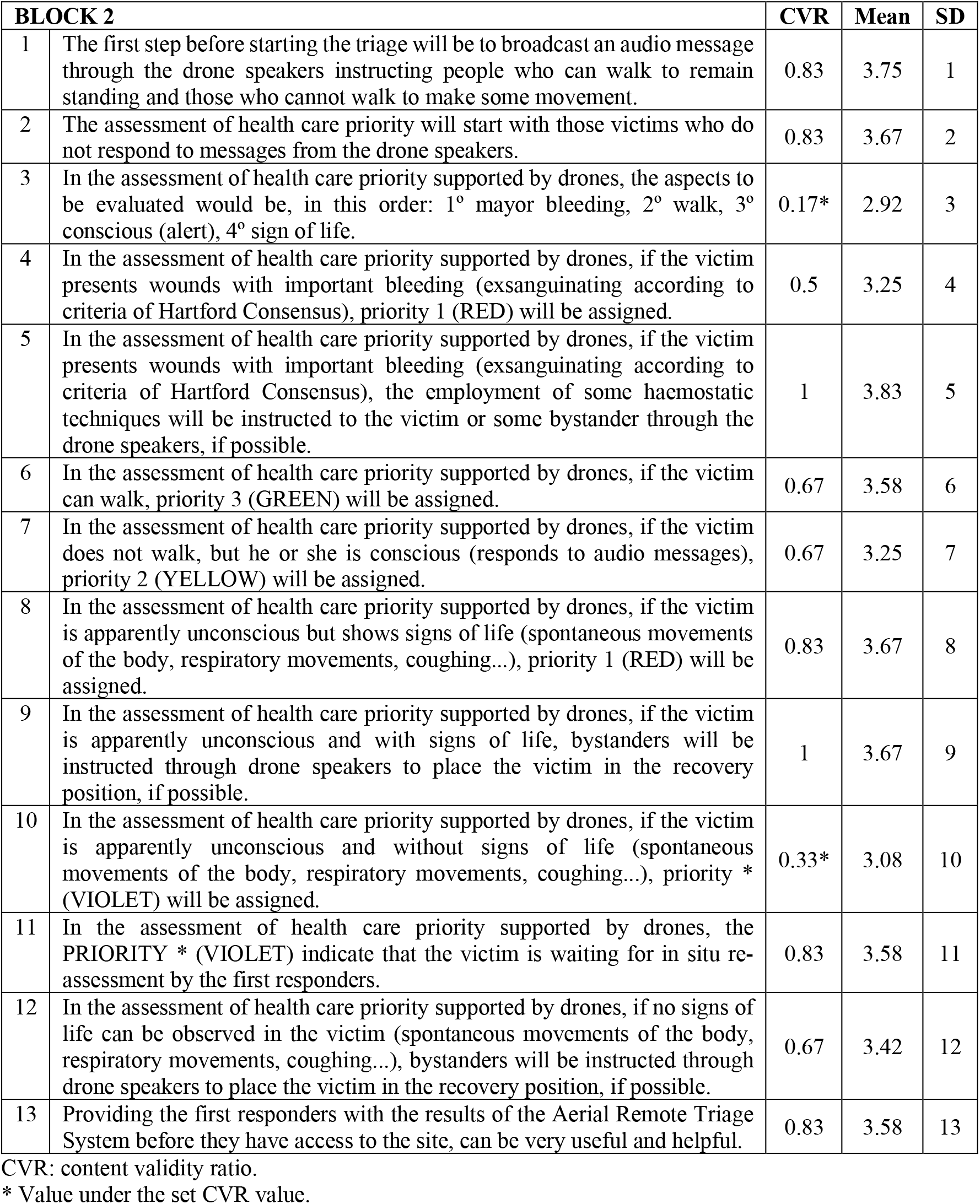
Content evaluation of statements about the use of the drones for triage.

We include in Table 4 some of the experts’ comments grouped into different categories in order to understand the experts’ consensus.

**Table 4.** Experts comments regarding the use of drones in health emergencies grouped by topic.

| **Use of drones in health emergencies** |  |  |
| --- | --- | --- |
| 1. Use of drones in complex health emergency scenarios with no physical access | → For the evaluation of the incident.<br>→ For better management of the emergency. | ○ "The information collected ... is very relevant."<br>○ "... to introduce a drone to obtain images, ... is vital."<br>○ "Whenever there is a foreseeable risk on the scene we should opt for drones or robots that allow us to explore the situation without risk to human lives"<br>○ "...the drones can provide information about the victims in a safe and agile manner..." |
| 2. Usefulness of instructions broadcast through the drone's loudspeakers | → To provide medical support.<br>→ To communicate expressions of support to victims.<br>→ To request the collaboration of bystanders in the realization of life-saving interventions.<br>→ To indicate measures of self-protection to the victims. | ○ "A drone can provide emotional support and inform victims that emergency services are approaching and looking out for them."<br>○ "It can be an excellent alternative means of support to the traditional one, for example by indicating how to apply a tourniquet, or how to move a victim in a safe side position or to perform basic life-saving interventions."<br>○ "Yes, you can help by giving instructions to a person who may not have any information about where they are." |
| 3. Usefulness of the drone for the carriage of medical material |  | ○ "Yes, it could carry insulin in pens for diabetics, for example, or glucagon, or even some antidote for poisoning or chemical accidents, knowing for sure the agent producing the poisoning." |
| 4. Usefulness in assessing victims and prioritizing their care |  | ○ "Yes, having microphones and emergency personnel managing the incident from the transmitter side of the loudspeaker would be a perfectly valid and useful procedure." |
| **Aerial Remote Triage System** |  |  |
| 1. Previous step, broadcast a message (audio) through the drone | → To identify victims who can walk or move.<br>→ To begin prioritizing unresponsive victims. | ○ "In the message I would first add that those who understand the message should make a gesture (e.g. raise a hand)."<br>"I would do it this way, to assess whether they are conscious or present exitus, or life-threatening difficulties, are the red, black or grey victims and we have to differentiate them in this way. I think it's appropriate to do it this way." |
| 2. Sequence of aspects to be evaluated: 1° mayor bleeding, 2° walk, 3° conscious (alert), 4° sign of life | → To assign priority in the remote evaluation process. | ○ "This is the pattern of action according to the triage START."<br>"Those who need life-saving interventions first so they could be addressed." |
| 3. Victims with bleeding injury (Hartford Consensus) | → Priority 1 (RED) Drone loudspeaker indication of haemostasis techniques. | ○ "I think it's clear, given the mortality associated with a victim with significant bleeding that is exsanguinating." |
|  |  | ○ "Right. Ideally... the drone should have options to release a tourniquet and indicate its placement."<br>"Direct compression is easy enough to be performed by laymen." |
| 4. Victims with walking ability | → Priority 3 (GREEN) | "This is the pattern followed by different triage systems START, SHORT, Sieve, MRCC..." |
| 5. Victims with no walking ability but conscious | → Priority 2 (YELLOW) | "...yellow in reevaluation. The bad thing is that we have no option to easily assess the neurological one since it can be a yellow that is really red because of ECT or with neurological deficits." |
| 6. Unconscious victims with signs of life (presence of any spontaneous body movement, breathing movements, coughing...) | → Priority 1 (RED)<br>→ Recovery position | ○ "Yes, I agree, it's the data that indicates us he's at least breathing and has signs of life."<br>○ "This would be ideal."<br>"We have to be very careful here as the victim may have injuries..." |
| 7. Victims apparently unconscious with no signs of life | → Priority * (VIOLET)<br>→ Victim waiting to be reassessed in situ by the first responders<br>Recovery position | ○ "Yes, I'm interested in reconfirming the victim's condition in person, for confirmation of the expected diagnosis."<br>○ "I agree when violet is an unevaluated patient"<br>"As long as it doesn't compromise the bystander in becoming another victim I see fit." |
| 8. Communication to the first responders of the findings of the Aerial Remote Triage System | → Usefulness of the information collected through the drones | ○ "Yes because it speeds up the organization of the team and the discrimination of attention to the victims who need it first."<br>○ "It can be of great help"<br>"Information is the most important thing in a catastrophe... To my mind, that wouldn't be the best thing... the best thing is for the drone to position the victims with its triage and take a picture." |

In relation to the statements 1 and 2 that point to the usefulness of the use of drones in complex health emergencies or in areas of difficult access for an initial assessment of the incident and an evaluation of the scenario, there is a high degree of agreement. The experts have mentioned the valuable possibility of gathering the necessary information in a health emergency situation by means of aerial images when there is no way to access the site. Especially, in radiation risk situations in nuclear incidents, chemical spills or suicide attacks, drones can help by avoiding exposure of people and identifying the characteristics and magnitude of the incident.

In statement number 3, related to the collection of relevant information for clinical assessment through drones as an alternative when ground access is not feasible, there was also a high agreement. Most participants referred to the safety and speed of the drone in providing information on the victims and valued its presence as an alternative when access to the scenario is not possible, such as in a flood. In other cases, doubts have been expressed about being able to make this clinical assessment due to limitations of the device, the legal restrictions on the use of the drone in some countries, or the difficulty of assessing the condition of the victim without being in situ.

According to statement number 4, related to the usefulness of the information provided by a drone in the decision-making process for emergency management, we found a high agreement among the experts. Experts have endorsed the idea that drones could be very useful in managing a health emergency since the information obtained through them about, for example, the presence of hazardous gases in a chemical release or the number of victims involved in the scenario and their location, is decisive in the moments following the incident. As one of the experts said: “More data and more information is always better”.

There is a consensus in statement 5 regarding the use of the drone’s loudspeakers to provide remote medical support to victims. It should be noted that several experts point out that it is possible to give instructions for compression of exsanguinating bleeding, to perform basic life support or to place the unconscious person in the recovery position. There were also experts who were concerned about one limitation, the drone do not allow the victims to communicate with the rescuers, so it is proposed the use of walkie talkies or mobile phones. Mention is also made of audio difficulties caused by the predictable noisy environment in typical collective emergency scenarios. In addition, they also suggest that this means of communication could be complemented with alternatives, such as laser light signals to indicate evacuation routes. Another limitation pointed out was the difficulty of offering emotional support.

The statements 6, was related to this last topic, the emotional support provided by drones, and the experts agree that it is possible and beneficial, despite the unidirectional nature of the communication. One of the experts proposed incorporating a screen in the drone that broadcasts the image of the person giving the instructions to make the process more humane.

In statement 7, a general consensus was reported regarding the capacity to “recruit”, through messages broadcast by the drone’s loudspeaker, those subjects involved in the incident who maintain their capabilities (bystanders) and who could help other more seriously injured people. One of the comments shows the value of using the drone to instruct victims to move to safer areas. Just one shortcoming was shown, the possibility that the confusion generated in a MCI, as well as the stress secondary to the impact, could make it difficult to follow orders. At the same time, some experts expressed doubts as to the authority or trust that a message issued by a “device” could generate.

Statement 8, which refers to the use of a loudspeaker on board a drone to give self-protection instructions to victims involved in a collective incident, the experts expressed the same level of consensus as in the previous statement and some of them expressed similar nuances to those already shown in the same previous section.

It was also reported a consensus in statement 9. This statement evaluated the possibility of issuing specific instructions to the bystanders to place victims in a recovery position or even apply haemostasis techniques if potentially bleeding wounds were detected. Some experts were reluctant about the capability of the bystanders to apply life-saving interventions.

The majority of the experts agreed on statement 10, which stated that it is possible that a drone sends useful medical equipment to the impact zone before the first responders’ arrival. It was proposed sending materials such as medication (antidote auto-injectors, analgesia), bleeding control kit or isothermal sheet. The experts also proposed sending other materials or devices, such as glucagon kit, automatic external defibrillators, as well as other types of materials, such as food.

Nonetheless, we found some disagreement in statement 11, referring to the individual assessment of victims using a drone, since some experts mentioned the difficulty of individualized assessment of victims in some cases, such as when the victim is in prone position, or the difficulty of assessing physiological parameters remotely. In this way, experts urge that triage be made as simple as possible.

Regarding the statements of the specific block of triage with drones, there is extensive agreement among experts on statement 1 regarding the broadcast of a message from the drone so that people who can walk stand up or those who cannot walk make some movement. The only aspect mentioned by the experts is the difficulty of understanding the message in noisy scenarios. There is also a broad consensus in statement 2 that priority should be given to victims who do not respond to the drone’s message. Only one expert mentions that perhaps those victims who do not respond are the least likely to survive and if a remote triage has been chosen the time until the victim can be attended to may be long, while other victims are neglected.

There was a disagreement on the sequence of assignment of priority in remote (statement 3), which was proposed to be first major bleeding, second walking, third conscious (awake) and fourth sign of life. The disagreement was due to the fact that some experts supported that this order is followed in other triage systems such as START, but others considered that it difficult to assess bleeding using images or it better to assess walking victims first so that they can move to a safe place. Nevertheless, the experts agreed on assigning priority 1 (red) to a victim with significant bleeding (statement 4), since the importance of applying haemostatic techniques to these victims was recognized (statement 5), but the difficulty of assessing bleeding remotely was mentioned. The possibility of sending tourniquets with the drone was introduced but the placement of these requires some training, so it was determined that the most appropriate technique that a bystander or the victim himself could use would be direct compression.

There was consensus among experts on the possibility, through the images provided by the drone, to classify wandering victims as priority 3 (green; statement 6). However, certain nuances are made regarding this category. The fact that they are able to walk does not indicate the absence of injuries. Hence the recommendation for continuous reassessment of these patients once they have been labelled green.

There is also an agreement with respect to the statement on the classification of a victim who cannot walk and is conscious as priority 2 (yellow; statement 7). Most of the comments refer to the limitations of the drone to accurately identify these victims, being important to discern if it is a yellow or red victim, and even if it is a green with walking problems. According to some experts, drones allow you to suspect a category but not rule it out. Others comment on the limitations of an aerial triage compared to standard triages, since with the former you can only count, evaluate and send messages.

On statement 8, which classifies an unconscious victim with signs of life as priority 1 (red), there is great agreement among experts. Some of the comments of the experts refer to the difficulty to detect, with the images provided by the drone, the presence of signs of life, among them respiratory movements, something complicated to achieve even being in contact with the victim. For others, it is simply a difficult challenge to achieve.

Although there is a high degree of agreement among experts on indicating a recovery position (statement 9), some of consider it relevance in this type of situation. They state that in the event of even the slightest suspicion of a spinal injury, this intervention should be avoided and the patient should not be mobilised.

In relation to statements 10 and 11 referring to the designation of the new violet colour used in this type of remote triage, we found different scores on the degree of agreement. Thus, there is some disagreement among the experts on the use of the colour violet to designate an apparently unconscious victim who does not show respiratory movements or some body movement (CVR=0.33), and they indicate the use of other already standardized colours such as black, red or blue. However, they indicate the use of the colour violet for those patients who need to be revaluated (CVR=0.83).

In statement 12, although there is agreement among the experts in the case of the use of life-saving interventions by some bystander, such as the placement of recovery position of those victims who remotely have not observed signs of life, they mentioned to the need to be cautious in carrying out such interventions in the face of the possibility that the victim presents vertebral injuries or injuries incompatible with life.

In statement 13, there is general agreement among the experts of an organized communication of the results of aerial remote triage with drones to the first responders. They even point out the additional advantage of determining the position of each of the victims previously classified in each of the categories, so that the deployment and use of resources (especially time) would be faster and more efficient.

Taking into account all the experts’ comments and the consensus reached, the Aerial Remote Triage System (ARTS) was built up and it is shown in Fig 1.

**Figure 1.**
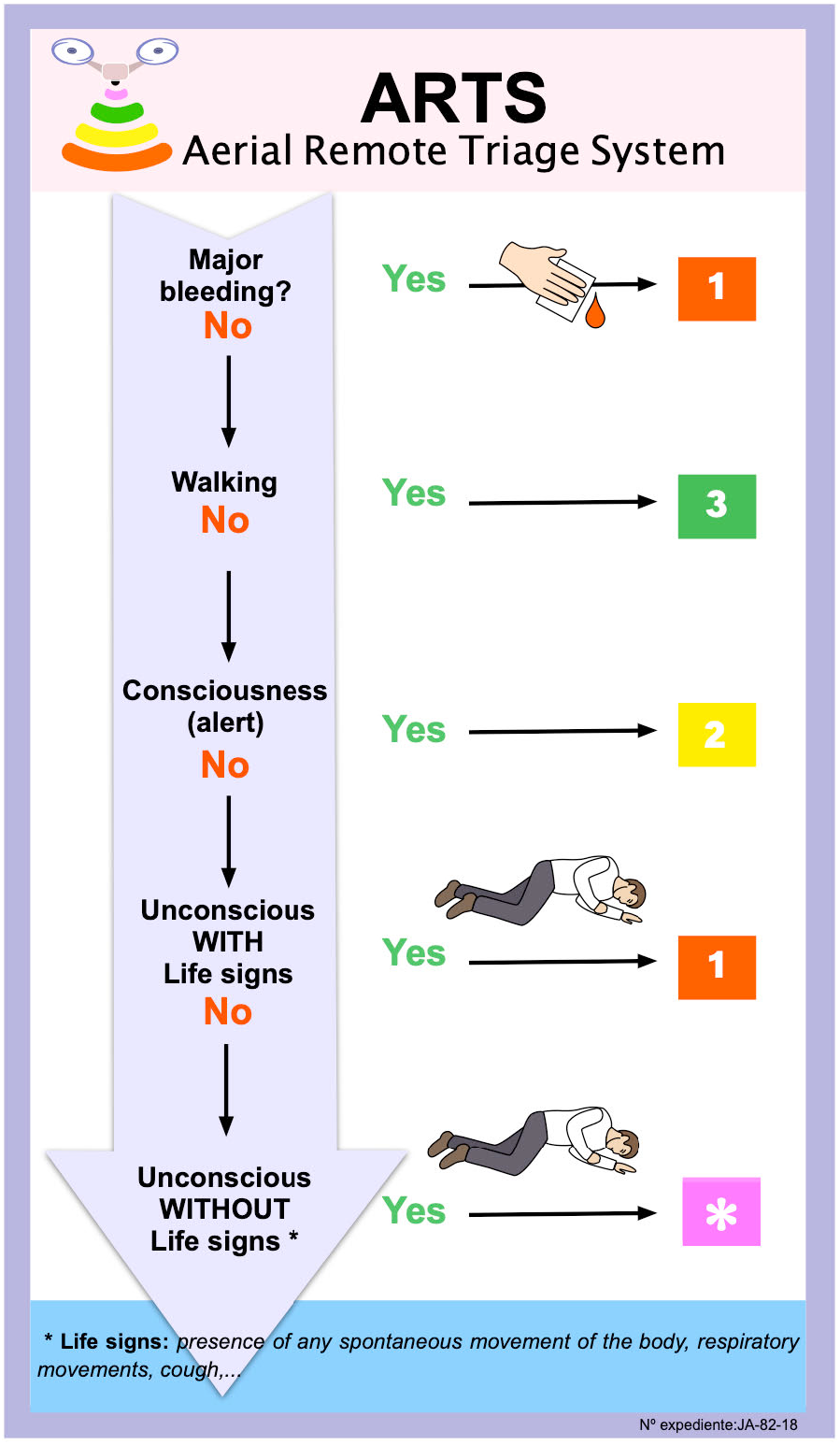
Aerial Remote Triage System.

## Discussion

The aim of our study was to develop the ARTS. The results of qualitative and quantitative analyses conducted through a panel survey demonstrate the utility of the ARTS in complex health emergencies. We discuss suggestions based on the consensus of the experts

Using drones for the assessment of complex health emergencies can be useful in enabling earlier assessment of the event by emergency teams when access to the incident site is difficult, or there is a risk to rescuers [3]. With an initial exploration flight, it is possible to determine the type of incident, its extension, the existence of added dangers, the number of victims, their distribution on the site and the possible access and evacuation routes [8, 22]. The use of drones has been shown to be accurate and safe in the identification of hazards in scene assessment as indicated in the study by Jain et al. [23] which compared the identification of seven hazards using unmanned aerial vehicles versus standard procedure. In both cases the hazards were identified at 100%. This makes it possible to implement the action plan earlier and thus improve incident management [13]. However, it is necessary to consider the recommendation of the Committee on Guidance for Establishing Crisis Standards of Care for Use in Disaster Situations, there is an obligation to make a prudent assessment in the allocation of available resources and to avoid situations where health care resources are diverted from the emergency to the deployment of drones without well defined, reasonable and informed objectives [24].

Previous studies have used voice commands to interact with victims in a MCI [15, 25-27]. Holgerson et al. [27] shown some strategies in order to overcome communication difficulties at the scene of a MCI, they include the reservation of priority lines in the mobile phone network and the use of loudspeakers to inform the crowds. A study by Jain et al. [4], in simulated MCI scenarios, showed that victims were able to follow the directions given to them by the first responders through a loudspeaker incorporated in a drone. Other study by Sanfridsson et al. [28] addressed the delivery of Automated External Defibrillators (AEDs) using drones in simulated out-of-hospital cardiorespiratory arrest scenarios. The results of the study showed that bystanders perceived less difficulty in interacting with the drone than they did in interacting with their own mobile phone in speakerphone mode or with the defibrillator itself. Carter et al [29] analysed the application of collective psychology to develop recommendations for the management of mass decontamination in nuclear, radiological, biological and chemical incidents. The authors concluded that the communication strategies that subjects perceived as most effective were those that included health-centered explanations, information about the actions that responders were taking, and sufficient practical information. Loudspeaker were used by the first responders, due to the nature of the scenario, and the victims could implement the recommended actions to decontaminate themselves.

The Hartford Consensus [30] established in 2017 the figure of the bystander as the subject who witnesses the incident but maintains his or her ability to help others who are injured. Besides, the Tactical Emergency Causality Care (TECC) Committee [30] indicate the performance of remote medical assessment techniques as well as giving indications to the injured to protect themselves, compress bleeding injuries or adopt recovery positions. It is necessary to clarify that, contrary to the traditional triage systems, in the ARTS we propose that the injured should not be asked to leave the area, since this would distance the recruited bystanders from the victims who could benefit from their help; as long as this does not pose a risk to these people.

The literature shows different studies that establish the performance of life-saving interventions during out-of-hospital triage in collective incidents. Lerner et al. [15] includes as intervention the control of exsanguinating haemorrhages, open the airway (recovery position) and the use of antidote-type medication in an auto-injector format. The recovery position can improve the overall prognosis of victims [31, 32]. The European Resuscitation Council Guidelines [33] refers to the performance of this intervention in all unconscious patients with spontaneous breathing, recommending caution in those victims with suspected spinal trauma. Studies comparing the recovery position with the HAINES (High Arm IN Endangered Spine) technique [34] on cadavers have not been able to demonstrate evidence on which is the best version of the recovery position in patients with cervical injuries [35]. The systematic review conducted by Hyldmo et al. [36], provides evidence that unconscious victims with trauma should be placed in the recovery position. Mesar et al. [37] proposed to send tourniquets, dressings or painkillers.

The experts shown some disagreement with respect to individual remote victim assessment, but we consider this point to be essential, since, despite being difficult, Sibley et al. [13] showed that 82% of health professionals involved in triage in a multiple-victim accident, adequately classified 12 of the 15 victims using the SALT method. In addition, Abrahamsen et al. [7] showed that a drone could identify the number of people involved in an MCI, their general condition, the presence of respiratory movements and the state of consciousness after simulations in different types of accidents. Regarding this, some experts mentioned the impossibility of determining physiological parameters of victims in prone position, but a recent study [38] has shown that it is possible to determine whether a person is alive or dead from the processing of images captured by a drone, since the cardiorespiratory movements of victims in the supine, prone or lateral position are evaluated. However, the objective of this work is limited to the development of a simple remote triage system, which does not require special skills to determine physiological parameters and is adapted for remote use, avoiding any errors such as those that occurred when using the SALT method in remote triage in the previously mentioned study [13].

Moving on to the proposed ARTS, agreement was reached regarding the emission of a message with the drone for a first classification of victims who can walk or at least make some movement, since this is a usual process in different triage systems and included in the Hartford-TECC Compendium [30]. The study by Jain et al. [4] showed that a drone is an equally effective alternative to a team of rescuers for an initial classification of victims by issuing a message to evacuate victims who are able to walk. Thus, once the less serious victims have been detected, since they at least maintain some ventilation and hemodynamic capacities, it is possible to begin to assess the victims who have not responded to the message and may be more likely to benefit from life-saving interventions applied by bystanders [31].

Despite some disagreement, the sequence of aspects to be assessed is finally accepted, as it is included in various triage guides [30, 39]. Furthermore, this has been the sequence used in the development of a triage system for laymen, non-citizen AID [40]. According to the statements related to the prioritization of victims with drones, there is generally great agreement among all experts. Although some experts considered that it should first be assessed whether victims can walk to classify them as green and that they can help other victims, one of the principles of triage is “fidelity”, based on which the benefit of people who need more help at the time of triage is considered preferable to the benefit of other victims [14]. In addition, it is widely known that a victim with significant bleeding can reach a state of shock and die of bleeding within minutes [41] and interventions such as direct wound compression could save his or her life [42], so it has been considered important to assess this aspect first.

Regarding the victims classified as priority 1 or green, it is necessary to re-evaluate and monitor them continuously once they have been identified. This measure is something that all traditional triage systems include within their methodologies and is a basic principle of this procedure [15, 31, 43].

In victims classified as priority 2 or yellow, given the difficulty of identifying respiration and its quality at a distance, it is proposed that a victim who is conscious is considered to be breathing and has a sufficient heartbeat to maintain cerebral perfusion, something that most terrestrial triage systems reflect in their procedures [43]. In this statement the experts express their doubts about whether the drone is capable of safely identifying this type of victim, since they may be in a worse or even better clinical situation than the one assigned. For some years now, some researchers [44, 45] have used techniques such as photoplethysmography and movement magnification, to obtain the heart and respiratory rate remotely with the help of images provided from a drone. Although some problems have to be solved, they show us the possibilities of drones.

For victims classified as priority 3 or red, it is proposed that any apparently unconscious victim with signs of life be labelled as such. Given the difficulty of obtaining a heartbeat remotely, the presence of respiratory movements, spontaneous body movements, coughing, …are taken as signs of life [33]. The comments of the experts refer mainly to this difficulty of drones to detect signs of life. In this regard, we must again recall the work of Al-Naji et al. [44] in which they obtain vital signs remotely through different imaging techniques and the study of Yamazaki et al. [46] in which they manage to detect victims under the rubble by means of a two-way audio system, capable of capturing the voice response and determining where it is located within the scene.

The use of standardized colours, universally accepted, is oriented to indicate the life expectancy of the victims, in which each of these assigned categories shows the time of action, as well as the therapeutic effort to be made [31]. Although the main colours used are red, yellow, green and black, there are other triage systems such as Homeboush that add other colours. Thus, they use white to designate injured people who have a pulse but are not breathing (in a state of death). These patients have more severe injuries (compared to those designated as red), but because they have signs of life they cannot be placed in the black group. In the Triage Early Warning Score model, the orange colour is used [14]. In the case of the triage algorithm presented in this study, the colour VIOLET was chosen because it is impossible to remotely confirm whether a patient who is unconscious and without signs of life is dead or not. The use of this colour indicates, on the one hand, the extreme seriousness of the victim awaiting confirmation of his condition by the first responders when they arrive at the scene and, on the other hand, as a way of differentiating it from the red category.

In view of the benefits of the use of drones in medicine, it is necessary to develop regulations at the global level for their correct use [47], taking into account the places and situations where their use would be allowed by doing a risk-benefit analysis [5, 7]. Among the regulations adopted in some countries, we can find the necessity to obtain a certificate to pilot the drone or the use of drones only during daylight hours, with the drone in the line of sight and in controlled airspaces [48]. It would also be necessary to control intrinsic factors of the drone such as flight speed, payload, characteristics and sensors that a medicalized drone must possess; as well as extrinsic factors, such as the weather conditions in which it can fly [49-52]. At present the main priority is to carry out more research in real situations since most of them have been carried out in simulated situations.

## Conclusions

The ARTS is valid in complex health emergencies when it is difficult or impossible an immediate access to the scene due physical, chemical or biological risks. It can be useful to know vital information about the emergencies. The system assesses first major bleeding, second walking, third conscious (awake) and fourth sign of life; and classify the victims inside priority categories: priority 1 (red), priority 2 (yellow), priority 3 (green) and priority * (violet). Besides, the system includes the possibility to indicate save-living interventions to victims and bystanders, like the compression of bleeding injuries or the recovery position.

## Data Availability

Data would be provided upon request

https://cuidsalud.com/en/portfolio/drones-emergencies/

## Acknowledgments

The authors would like to thank the experts who help in the process of developing the ARTS.

